# Healthy adults possess cross-reactive neuraminidase inhibition antibodies to an A(H5N1) clade 2.3.4.4b avian influenza virus A/Black Faced Spoonbill/Hong Kong/AFCD-HKU-22-21429-01012/2022

**DOI:** 10.1101/2023.06.23.23291839

**Authors:** Pavithra Daulagala, Samuel M. S. Cheng, Alex W. H. Chin, Leo L. H. Luk, Kathy Leung, Joseph T. Wu, Leo L. M. Poon, Malik Peiris, Hui-Ling Yen

## Abstract

Hemagglutination inhibition (HI) and neuraminidase inhibition (NI) antibodies to a clade 2.3.4.4b A(H5N1) highly pathogenic avian influenza virus were measured in 63 age-stratified healthy adults in Hong Kong. No HI antibody was detected; 61 subjects had detectable NI antibodies to A(H5N1). NI titers to A(H5N1) and A(H1N1)pdm09 viruses were correlated.

## Text

The A/Goose/Guangdong/1/1996-like (GsGd-like) A(H5N1) highly pathogenic avian influenza viruses were first identified in 1996 and have continuously evolved into distinct HA clades that are antigenically different. Prior to 2005, the GsGd-like virus mainly circulated in Asia among domestic poultry (including land-based poultry and domestic waterfowls). Spill-over infections from domestic poultry to wild migratory birds have facilitated long-distance spread into different continents (1, 2). In 2005, the clade 2.2 viruses spread from China to Europe, the Middle East, and Africa. In 2014-2015, the clade 2.3.4.4c viruses with new NA proteins acquired through genetic reassortment spread from Asia to Europe and North America. Since 2016, the clade 2.3.4.4b viruses have continued to cause outbreaks in domestic poultry in Europe (3) and become enzootic among wild birds in 2021 (4), and further spread to North America and South America in 2021-2022. With expanded genetic diversity and geographic distribution, spill-over events into numerous mammalian species and sporadic human infections were also documented (5). While the HPAI (H5N1) virus has not yet achieved efficient transmissibility in humans, the current epidemiology of A(H5N1) 2.3.4.4b lineage raises pandemic concern.

Population immunity to an emerging influenza virus is one of the key parameters considered while assessing its pandemic risk (6, 7). Neutralizing antibodies targeting the receptor binding domain of HA and antibodies that inhibit NA activity have been shown to correlate with protection against influenza infection (8, 9). Here, we aimed to evaluate if the general population possess cross-reactive antibody responses to the A(H5N1) through prior exposures to seasonal influenza viruses. Sera from healthy blood donors (N=63) aged 18-73 years (born between 1947-2002) collected from April 2020 – August 2020 in Hong Kong (HKU/HA HKW Institutional Review Board approval #UW-132) were used to determine cross-reactive HI antibodies and NI antibodies against a HPAI (H5N1) clade 2.3.4.4b virus A/Black Faced Spoonbill/Hong Kong/AFCD-HKU-22-21429-01012/2022 (Spoonbill/HK/22) isolated in November 2022. The HA protein of Spoonbill/HK/22 (GISAID: EPI2581007) shared 98.8% and 98.6% homology to the HA proteins of candidate vaccine viruses (CVVs) of clade 2.3.4.4b; A/chicken/Ghana/AVL-76321VIR7050-39/2021 and A/American wigeon/South Carolina/AH0195145/2021, respectively. Similarly, the NA protein of Spoonbill/HK/22 (GISAID: EPI2581009) shared a homology of 97.3% and 97.2% with the NA proteins of CVVs; A/chicken/Ghana/AVL-76321VIR7050-39/2021 and A/American wigeon/South Carolina/AH0195145/2021, respectively (10). As H5 and H1 belong to Group 1 HA, we also determined the HI and NI antibody responses towards A/California/04/2009 (California/09) A(H1N1)pdm09 virus, as exposure to A(H1N1)pdm09 may generate cross-reactive HI and NI antibodies to A(H5N1). HI and NI antibodies were determined using hemagglutination inhibition assay (11) and enzyme-linked lectin assay (12), respectively.

Among 63 healthy adults, 53/63 (84.1%) possessed detectable HI antibodies at ≥1:10 to California/09, with the highest HI response observed in the 10-19 years old age group. However, none showed detectable HI antibodies at ≥1:10 to Spoonbill/HK/22 (Figure 1). 57/63 (90.5%) healthy subjects showed detectable NI antibodies at ≥1:10 against California/09, with the highest NI response observed in the 10-19 years old age group. Interestingly, 61/63 (96.8%) healthy subjects possessed cross-reactive NI response at ≥1:10 and 36/63 (57.1%) had NI titres of ≥1:40 to Spoonbill/HK/22. The NI titers against California/09 and Spoonbill/HK/22 were highly correlated (r=0.8349, Spearman’s correlation, p <0.001). Specifically, 57 (90.5%) and 32 (50.8%) subjects had NI antibodies to both viruses at titers ≥ 1:10 and ≥1:40, respectively.

**Figure 1.**
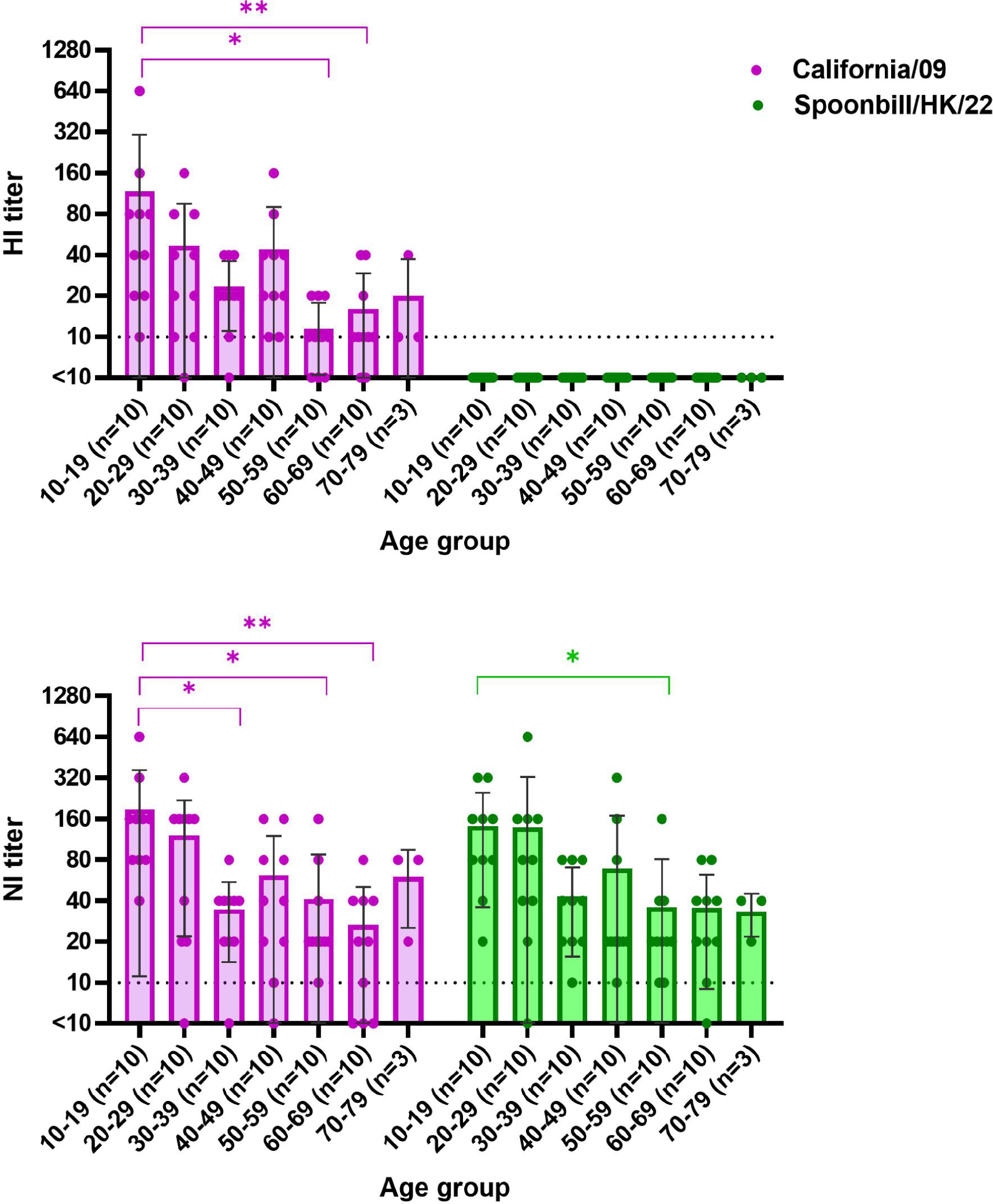
HI and NI antibody responses against California/09 and H5N1 with age in healthy adults. A scatter graph with a bar showing the HI and NI antibody response measured against California/09 (H1N1) and Spoonbill/HK/22 (H5N1) in different age groups with an x-axis showing the age group and y-axis showing the HI and NI titers measured. The sample size of each age group is shown within the brackets (n=x). The HI and NI titers measured against California/09 are shown in purple, and the HI and NI titers measured against Spoonbill/HK/22 are shown in green. Each dot in an age group represents a HI or NI titer measured for an individual. The bar generated for the age group represents the mean with standard deviation shown in the black line. Individuals showing HI and NI titer below the detection limit is shown as <1:10. The HI and NI titers across different age groups for each virus was compared using Kruskal Wallis test and Dunn’s multiple comparison test. The ^*^ p<0.033, ^* *^p<0.002, ^* * *^ p<0.001.

To delineate the potential exposure history in healthy adults that lead to cross-reactive NI response to Spoonbill/HK/22, archival ferret anti-sera raised against A(H1N1) viruses and A(H1N1)pdm09 (California/09) were used to determine NI responses against the homologous virus and Spoonbill/HK/22 (Table 1). Ferret anti-sera raised against A(H1N1) viruses circulated in 1977-2007 showed no cross-reactive NI response to Spoonbill/HK/22, despite of detectable NI titers at 1:320 to 1:1280 against the homologous viruses. Ferret anti-sera raised against California/09 showed a homologous NI titer of 1:2560 and cross-reactive NI titres to Spoonbill/HK/22 at 1:320 to 1:640. The NA proteins of Spoonbill/HK/22 and California/09 differed by 53 amino acids (88.1% homology), while the NA proteins of Spoonbill/HK/22 and seasonal A(H1N1) viruses differed by 68 – 76 amino acids (84.1% to 84.6% homology) (Table 2). More amino acid substitutions were observed in the NA head domain of A/Brisbane/59/2007 (Brisbane/07) (n=54 amino acid differences) than California/09 H1N1 viruses (n=34 amino acid differences), in comparison to Spoonbill/HK/22 (Figure 2 and Table 2).

**Table 1.** NI antibody response in post-infection ferret anti-sera against NA of Spoonbill/HK/22 and California/09.

| Post-infection<br>ferret anti-sera | NI antibody titer measured using A(H6N1)<br>viruses |  |  |
| --- | --- | --- | --- |
|  | Homologous<br>virus<br>A(H1N1) | California/09<br>A(H1N1) | Spoonbill/HK/22<br>A(H5N1) |
| A/USSR/90/1977 | 320 | <10 | <10 |
| A/Chile/01/1983 | 320 | <10 | <10 |
| A/Singapore/06/1986 | 320 | <10 | <10 |
| A/Texas/36/1991 | 320 | <10 | <10 |
| A/Brisbane/59/2007 | 1280 | <10 | <10 |
| A/California/04/2009_pdm09 | 2560 | 2560 | 320-640* |

\*NI titer range detected in n=6 ferret anti-sera

**Table 2.** Number of amino acid differences in the NA proteins of A(H1N1) and A(H1N1)pdm09 viruses in comparison to A(H5N1)

| A(H1N1) strain | Amino acid differences compared to Spoonbill/HK/22 | Homology (%) |
| --- | --- | --- |
| A/USSR/90/1977 | 70 | 84.08 |
| A/Chile/01/1983 | 68 | 84.58 |
| A/Singapore/06/1986 | 68 | 84.58 |
| A/Texas/36/1991 | 69 | 84.33 |
| A/Brisbane/59/2007 | 76 | 83.83 |
| A/California/04/2009_pdm09 | 53 | 88.00 |

**Figure 2.**
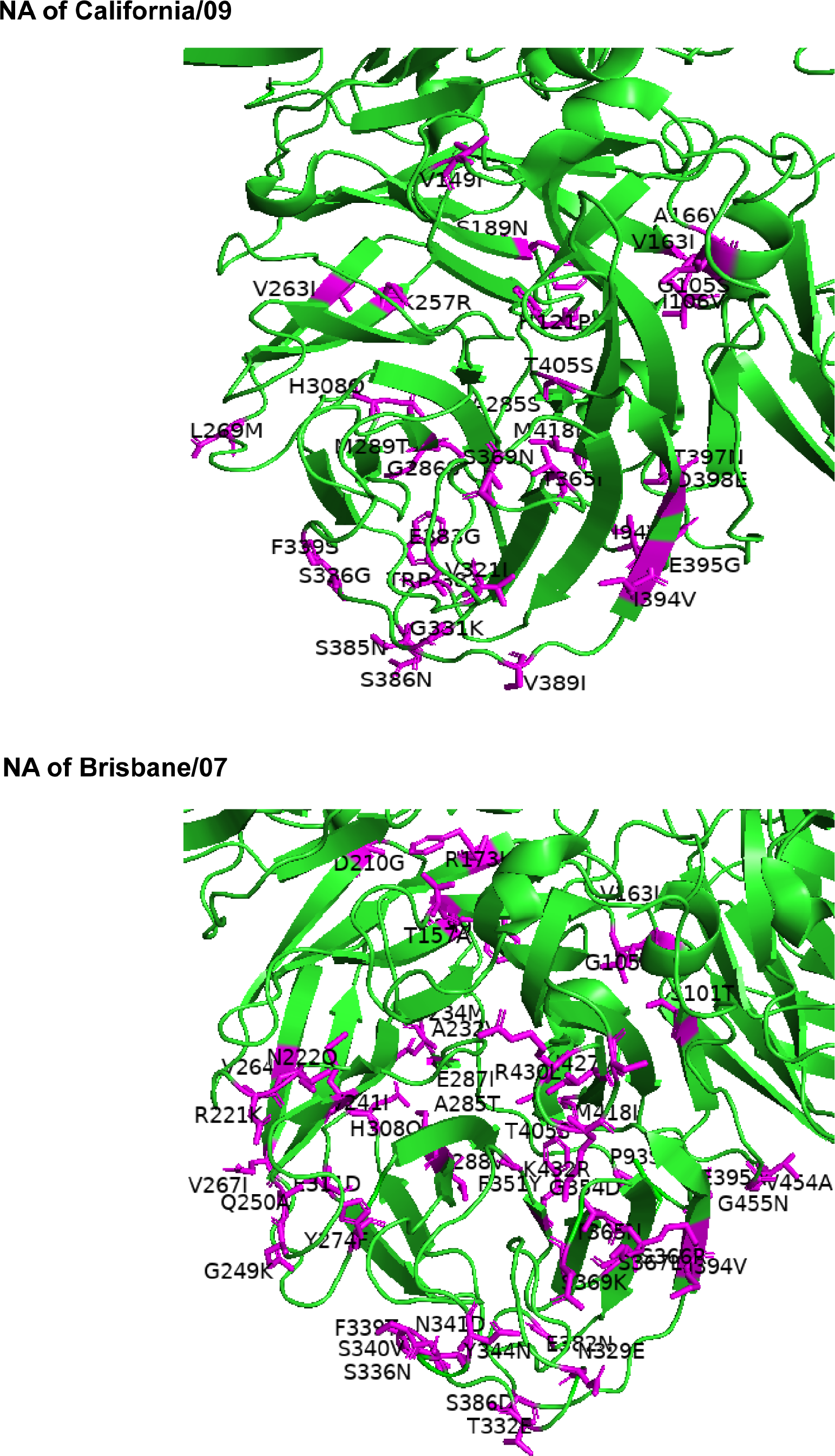
Amino acid substitutions on NA proteins of California/09 and Brisbane/07 in comparison to the NA of 2.3.4.4b A(H5N1) virus, Spoonbill/HK/22. The positions of the amino acid differences in the head region of NA are shown on the NA tetrameric structure of A/Vietnam/1203/2004 (PDB 2HU0) generated using the software PyMOL. The NA monomers are indicated in green and the amino acid substitutions are labelled in purple.

## Conclusion

We detected cross-reactive NI antibodies among healthy adults against a clade 2.3.4.4b A(H5N1) virus, Spoonbill/HK/22, a virus genetically closely related to A(H5N1) viruses recently circulating in Europe and the Americas. The NI response to A(H5N1) was highly correlated with the NI response to the A(H1N1)pdm09 virus. Our results confirm and extend the findings from a recent study on detecting cross-reactive NI antibody responses to clade 2.3.4.4b A(H5N1) virus in healthy blood donors (13). Using archival ferret anti-sera raised against seasonal A(H1N1) and A(H1N1)pdm09 influenza viruses, cross-reactive NI response to A(H5N1) was only detected in ferret sera infected with A(H1N1)pdm09, but not seasonal A(H1N1) viruses circulated in 1977-2007. The A(H1N1)pdm09 virus derived its NA protein from the avian-origin Eurasian-avian swine viruses (14) and appeared antigenically more closely related to the N1 of recent clade 2.3.4.4b H5N1 viruses and may thus provide partial protection against H5N1 HPAIV infection. Anti-NA antibodies have been identified to correlate with protection, specifically to protect against infection, reduce symptoms, and shorten the duration of viral shedding (9). HI titer of ≥1:40 has long been established as a surrogate for protection against influenza virus infections, allowing for estimating the impact of cross-reactive HI antibody titers in age-stratified human sera on reducing the reproduction number R0 of a novel zoonotic virus that may acquire transmissibility in humans (15). However, levels of NI antibodies that correlate with protection have not been clearly defined and it is therefore not possible to assess the impact of the observed cross-reactive NI antibody to A(H5N1) at the population level.

Taken together, high titers of cross-reactive NI antibodies against NA of HPAI (H5N1) clade 2.3.4.4b viruses are detected in human sera, although there are no cross-reactive HI antibodies. This suggests that NI antibodies generated against conserved epitopes across human and avian N1 NA may partially protect humans against A(H5N1) infection or modulate disease severity.

### Biographical sketch

Dr. Daulagala is a researcher at the School of Public Health, University of Hong Kong. Her research interest is to understand the role of anti-NA antibodies against influenza infections.

## Data Availability

All data produced in the present study are available upon reasonable request to the authors

## Acknowledgements

The authors thank Dr. Christopher J. Brackman from the Agriculture Fisheries and Conservation Department (AFCD), Government of the Hong Kong Special Administrative Region (SAR), China, for sharing the A(H5N1) virus. This study was supported by RGC Theme-based Research Schemes (T11-712/19-N), Hong Kong SAR, China.

## References

1. Caliendo V, Lewis NS, Pohlmann A, Baillie SR, Banyard AC, Beer M, et al. Transatlantic spread of highly pathogenic avian influenza H5N1 by wild birds from Europe to North America in 2021. Sci Rep. 2022;12(1):11729. Epub 20220711. doi: 10.1038/s41598-022-13447-z. PubMed PMID: 35821511; PubMed Central PMCID: PMC9276711.

2. Verhagen JH, Fouchier RAM, Lewis N. Highly Pathogenic Avian Influenza Viruses at the Wild-Domestic Bird Interface in Europe: Future Directions for Research and Surveillance. Viruses. 2021;13(2). Epub 20210130. doi: 10.3390/v13020212. PubMed PMID: 33573231; PubMed Central PMCID: PMC7912471.

3. European Food Safety Authority ECfDP, Control EURLfAI, Adlhoch C, Fusaro A, Gonzales JL, Kuiken T, et al. Avian influenza overview September -December 2022. EFSA J. 2023;21(1):e07786. Epub 20230119. doi: 10.2903/j.efsa.2023.7786. PubMed PMID: 36698491; PubMed Central PMCID: PMC9851911.

4. Pohlmann A, King J, Fusaro A, Zecchin B, Banyard AC, Brown IH, et al. Has Epizootic Become Enzootic? Evidence for a Fundamental Change in the Infection Dynamics of Highly Pathogenic Avian Influenza in Europe, 2021. mBio. 2022;13(4):e0060922. Epub 20220621. doi: 10.1128/mbio.00609-22. PubMed PMID: 35726917; PubMed Central PMCID: PMC9426456.

5. WHO. Antigenic and genetic characteristics of zoonotic influenza A viruses and development of candidate vaccine viruses for pandemic preparedness: Geneva: World Health Organization.; 2022. Available from: https://cdn.who.int/media/docs/default-source/influenza/who-influenza-recommendations/vcmnorthern-hemisphere-recommendation-2022-2023/202203_zoonotic_vaccinevirusupdate.pdf.

6. Cox NJ, Trock SC, Burke SA. Pandemic preparedness and the Influenza Risk Assessment Tool (IRAT). Curr Top Microbiol Immunol. 2014;385:119–36. doi: 10.1007/82_2014_419. PubMed PMID: 25085014.

7. WHO. Tool for Influenza Pandemic Risk Assessment (TIPRA). 2 ed: Geneva: World Health Organization.; 2020.

8. Krammer F, Weir JP, Engelhardt O, Katz JM, Cox RJ. Meeting report and review: Immunological assays and correlates of protection for next-generation influenza vaccines. Influenza and Other Respiratory Viruses. 2020;14(2):237–43. doi: https://doi.org/10.1111/irv.12706.

9. Maier HE, Nachbagauer R, Kuan G, Ng S, Lopez R, Sanchez N, et al. Pre-existing Antineuraminidase Antibodies Are Associated With Shortened Duration of Influenza A(H1N1)pdm Virus Shedding and Illness in Naturally Infected Adults. Clin Infect Dis. 2020;70(11):2290–7. doi: 10.1093/cid/ciz639. PubMed PMID: 31300819; PubMed Central PMCID: PMC7245146.

10. WHO. Summary of status of development and availability of A(H5N1) candidate vaccine viruses and potency testing reagents Geneva: World Health Organization2023. Available from: https://cdn.who.int/media/docs/default-source/influenza/cvvs/cvv-zoonotic-northern-hemipshere-2023-2024/h5n1_summary_a_h5n1_cvv_20230225.pdf?sfvrsn=cccc2a09_3&download=true.

11. WHO. Manual for the laboratory diagnosis and virological surveillance of influenza. Geneva: World Health Organization; 2011.

12. Couzens L, Gao J, Westgeest K, Sandbulte M, Lugovtsev V, Fouchier R, et al. An optimized enzyme-linked lectin assay to measure influenza A virus neuraminidase inhibition antibody titers in human sera. J Virol Methods. 2014;210:7–14. Epub 20140916. doi: 10.1016/j.jviromet.2014.09.003. PubMed PMID: 25233882.

13. Kandeil A, Patton C, Jones JC, Jeevan T, Harrington WN, Trifkovic S, et al. Rapid evolution of A(H5N1) influenza viruses after intercontinental spread to North America. Nat Commun. 2023;14(1):3082. Epub 20230529. doi: 10.1038/s41467-023-38415-7. PubMed PMID: 37248261; PubMed Central PMCID: PMC10227026.

14. Garten RJ, Davis CT, Russell CA, Shu B, Lindstrom S, Balish A, et al. Antigenic and genetic characteristics of swine-origin 2009 A(H1N1) influenza viruses circulating in humans. Science. 2009;325(5937):197–201. Epub 20090522. doi: 10.1126/science.1176225. PubMed PMID: 19465683; PubMed Central PMCID: PMC3250984.

15. Cheung JTL, Tsang TK, Yen HL, Perera R, Mok CKP, Lin YP, et al. Determining Existing Human Population Immunity as Part of Assessing Influenza Pandemic Risk. Emerg Infect Dis. 2022;28(5):977–85. doi: 10.3201/eid2805.211965. PubMed PMID: 35447069; PubMed Central PMCID: PMC9045452.

